# Comprehensive analyses reveal the impacts of vaccination status and physiological variables in early infection on viral persistence in COVID-19 patients: a retrospective single-center cohort study

**DOI:** 10.1101/2022.04.14.22273860

**Authors:** Xiangxiang Tian, Yifan Zhang, Wanhai Wang, Fang Fang, Wenhong Zhang, Yanmin Wan, Zhaoqin Zhu

**Affiliations:** Department of Laboratory Medicine, Shanghai Public Health Clinical Center, Fudan University, Shanghai, China; Department of Infectious Diseases, Shanghai Key Laboratory of Infectious Diseases and Biosafety Emergency Response, National Medical Center for Infectious Diseases, Huashan Hospital, Fudan University, Shanghai, China; Clinical Laboratory, The First Affiliated Hospital of Zhengzhou University, Key Laboratory of Laboratory Medicine of Henan Province, Zhengzhou, China; Shanghai Public Health Clinical Center, Fudan University, Shanghai, China; Department of radiology, Shanghai Public Health Clinical Center, Fudan University, Shanghai, China; State Key Laboratory of Genetic Engineering, School of Life Science, Fudan University, Shanghai, China

**Keywords:** COVID-19, inactivated vaccine, mRNA vaccine, laboratory variables, viral RNA shedding

## Abstract

**BACKGROUND:** Viral persistence is a crucial factor that influences the communicability of SARS-CoV-2 infection. However, the impacts of vaccination status and physiological variables on viral RNA shedding have not been adequately clarified.

**METHODS:** In this study, we retrospectively collected the clinical records of 377 hospitalized COVID-19 patients, which contained unvaccinated patients and patients received two doses of an inactivated vaccine or an mRNA vaccine. Firstly, we analyzed the impacts of vaccination on disease severity and viral RNA persistence. Next, to clarify the impacts of physiological variables on viral RNA shedding in COVID-19 patients, we retrieved 49 laboratory variables and analyzed their correlations with the duration of viral RNA shedding. Finally, we established a multivariate regression model to predict the duration of viral RNA shedding.

**RESULTS:** Our results showed that both inactivated and mRNA vaccines significantly reduced the rate of moderate cases, while the vaccine related shortening of viral RNA shedding were only observed in moderate patients. Correlation analysis showed that 10 significant laboratory variables were shared by the unvaccinated mild patients and mild patients inoculated with an inactivated vaccine, but not by the mild patients inoculated with an mRNA vaccine. Moreover, we demonstrated that a multivariate regression model established based on the variables correlating with viral persistence in unvaccinated mild patients could predict the duration of viral shedding for all groups of patients.

**CONCLUSIONS:** Vaccination contributed limitedly to the clearance viral RNA in COVID-19 patients. While, laboratory variables in early infection could predict the persistence of viral RNA.

## Introduction

Real world studies suggest that the implementing of vaccination have dramatically reduced the rates of SARS-CoV-2 infection and SARS-CoV-2 related hospitalization, admission to intensive care unit and death (1-3). In addition, retrospective estimations have also suggested that vaccines were effective at preventing the transmission of both the Alpha (4, 5) and the Delta variants (6). It is believed that the vaccine mediated reduction of transmission relies on the prevention of infection, while the effect of vaccines in preventing onward transmission after breakthrough infections has not been adequately clarified.

It is postulated that vaccines may help to restrain the onward transmission of SARS-CoV-2 (7) based on observations that COVID-19 vaccination can reduce viral loads in nasal mucosa (8-10). However, contradictory evidence showed that the viral loads of breakthrough infections after full vaccination were similar with those in unvaccinated individuals (11, 12). More recently, a preprint study carried out in a prison demonstrated that there was no significant difference in the duration of RT-PCR positivity and the kinetics of Ct values between fully vaccinated participants and those not fully vaccinated (13).

Given that the duration of virus shedding may have crucial impact on the onward transmission of SARS-CoV-2, in this study, we retrieved the clinical records of 377 hospitalized COVID-19 patients and analyzed the impacts of vaccination status and physiological variables in early infection on the duration of viral RNA shedding. Comparisons between fully vaccinated and unvaccinated patients showed that vaccination shortened the duration of viral RNA shedding among moderate COVID-19 patients, while showed no effect in constraining the duration of viral RNA shedding among mild COVID-19 patients. Furthermore, we showed that vaccination status impacted the association between individual physiological parameters and the duration of viral RNA shedding. Nonetheless, multivariate regression analyses showed that variables correlated with viral persistence in unvaccinated mild patients could predict the duration of viral RNA shedding for all groups of patients.

## Materials and methods

### Ethics statement

This study was approved by the Research Ethics Review Committee (Ethics Approval Number: YJ-2020-S080-02) of the Shanghai Public Health Clinical Center Affiliated to Fudan University.

### Study design and participants

377 COVID-19 patients hospitalized in Shanghai Public Health Clinical Center during the period from January 2020 to mid-January 2022 were included in this study. The diagnosis and clinical management of COVID-19 patients were conducted following the Diagnosis and Treatment Protocol for Novel Coronavirus Pneumonia released by National Health Commission & National Administration of Traditional Chinese Medicine (14). The demographic and COVID-19 vaccination information was depicted in Table 1. 49 laboratory variables detected at the immediate early stage of infection and the duration of viral RNA shedding were shown in Supplemental table 1. The duration of viral RNA shedding was defined as the period from the initial nasal swab RT-PCR positivity to the final nasal swab RT-PCR positivity. Patients with identified comorbidities were excluded from this study.

**Table 1.** Demographical characteristics of COVID-19 patients

|  | Inactivated vaccine<br>(N=165) | mRNA vaccine<br>(N=41) | Without<br>vaccination<br>(N=171) | P value |
| --- | --- | --- | --- | --- |
| **Gender (n)** |  |  |  |  |
| **Male** | 126 | 21 | 105 | 0.0011 |
| **Female** | 39 | 20 | 66 |  |
| **Age, Median (IQR)**<br>**(Years)** | 35.0 (25.0-44.0) | 30.0 (25.0-46.5) | 32.0 (24.0-44.0) | 0.5651 |
| **Time-since-vaccination**<br>**(Mean±SD) (Days)** | 175.5±90.83 | 146.0±63.63 | - | 0.1183 |

**Note:** All the vaccinated patients received two doses of either an inactivated vaccine or an mRNA vaccine. Time-since-vaccination is defined as the period from the second dose of vaccination to the diagnosis of COVID-19.

### SARS-CoV-2 viral RNA detection

Viral RNA extraction was performed using an automatic nucleic acid extractor (Bioperfectus technologies Co., LTD., Jiangsu, China). 2019-nCoV nucleic acid detection kit (Bioperfectus technologies Co., LTD., Jiangsu, China) and ABI 7500 real-time fluorescence quantitative PCR (qPCR) instrument (Thermo Fisher Technology Co., LTD., Shanghai, China) were used to detected SARS-CoV-2 genomic RNA. When the Ct values of both ORFlab and N gene of 2019-nCoV are ≤ 37 and the amplification curve is “S-shaped”, the result is defined as positive. When the Ct values of both genes are > 40 or undetermined, the result is defined as negative. When the Ct value is between 37∼40, the sample would be re-examined.

### Laboratory measurements

All the laboratory tests were carried out following manufacturers’ instructions in the department of laboratory medicine of Shanghai Public Health Clinical Center. All the tests had passed the ISO15189 accreditation. In this study, we retrospectively retrieved the values of 49 laboratory variables (Supplemental table 1) for each patient, which were measured at a median of 3 days post diagnosis.

### Statistical analysis

Quantitative data were examined for normality using the Shapiro–Wilk test before all downstream analyses except the Chi square test. Means for variables with a normal distribution were compared using the two-tailed parametric t-test and with the two-tailed non-parametric t-test when distributions of data departed from normality. Pearson correlation was used for correlation analyses of normally distributed data and Spearman correlation was used for analyses of non-normally distributed data. Multivariate regression analyses were performed for variables with statistical significance by the correlation analysis. P ≤ 0.05 was considered as statistically significant. All the statistical analyses were conducted using Graphpad Prism 9 (GraphPad Software, USA).

## Results

### Vaccines reduced the proportion of the moderate cases and shortened the duration of viral RNA shedding among moderate COVID-19 patients

In this study, we retrospectively retrieved the clinical records of 377 COVID-19 patients hospitalized in Shanghai Public Health Clinical Center during the period from January 2020 to mid-January 2022. Among these patients, 165 received two doses of an inactivated vaccine, 41 received two doses of an mRNA vaccine and 171 were not vaccinated (W/O vaccination) (Table 1). The median age was similar across all groups (P=0.5651), and the mean time-since-vaccination of patients inoculated with an inactivated vaccine was comparable with that of patients inoculated with an mRNA vaccine (P=0.1183) (Table 1). The gender composition of patients inoculated with an mRNA vaccine was significantly different from the other two groups, which contained more female (P = 0.0011) (Table 1), but it did not impact the duration of viral RNA shedding and the severity of disease (Supplemental figure 1).

Compared with unvaccinated patients, patients inoculated with either an inactivated vaccine (19.4% vs 35.7%, P=0.0002) or an mRNA vaccine (7.3% vs 35.7%, P=0.0001) significantly reduced the incidence of moderate cases (Figure 1A). More interestingly, we found that patients inoculated with an inactivated vaccine showed significantly shorter duration of viral RNA shedding in moderate patients compared with unvaccinated patients (17, IQR 12.25-19.5 vs 19, IQR 12-24.5, P=0.0382) and patients inoculated with an mRNA vaccine (17, IQR 12.25-19.5 vs 22.33 ± 4.163, P=0.0385) (Figure 1B).

**Figure 1.**
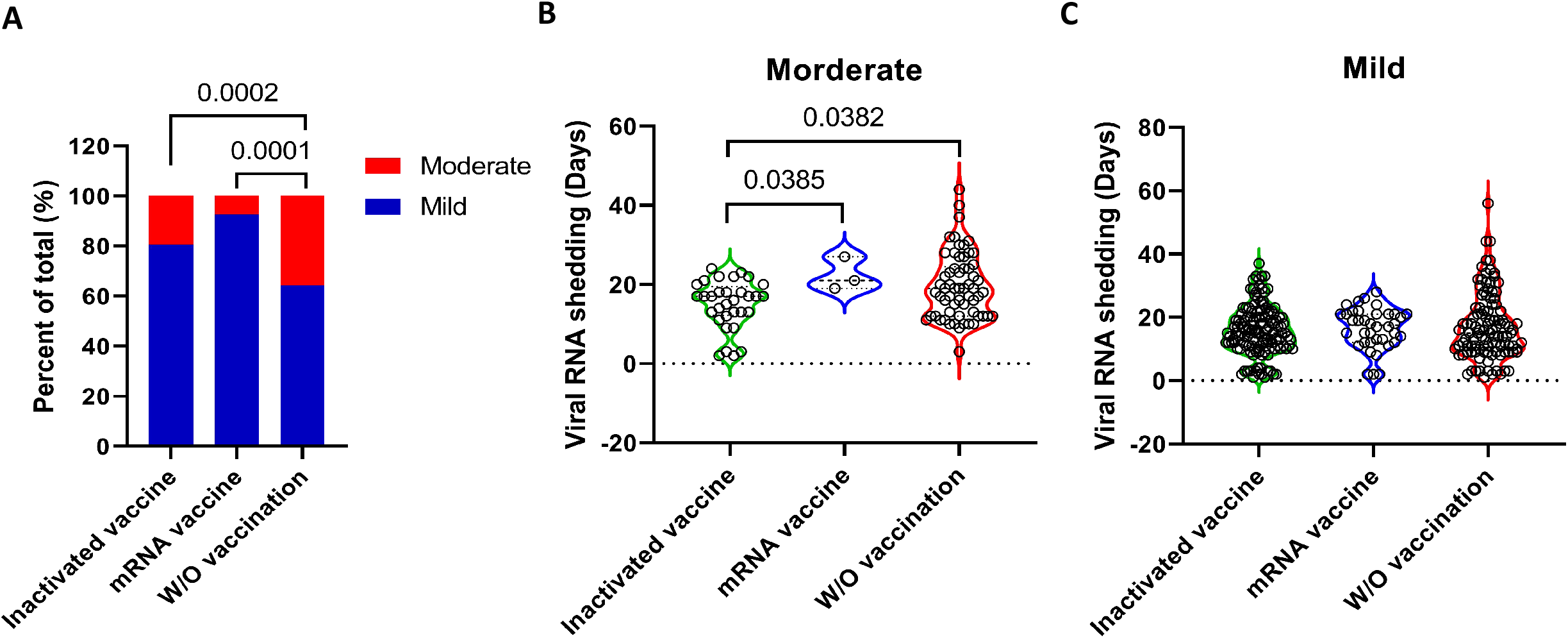
The impacts of vaccination on COVID-19 clinical manifestation and the duration of viral RNA shedding. (**A**) The proportions of mild and moderate COVID-19 cases were compared among unvaccinated patients and patients inoculated with an inactivated vaccine or an mRNA vaccine. (**B**) Comparisons of the duration of viral RNA shedding among moderate COVID-19 patients. (**C**) Comparisons of the duration of viral RNA shedding among mild COVID-19 patients. Statistical analyses were performed by the method of non-parametric t-test.

However, the vaccines showed no effect in constraining the duration of viral RNA shedding in mild COVID-19 patients (Figure 1C).

### Common variables that correlated with the duration of viral RNA shedding were found in unvaccinated mild patients and mild patients inoculated with an inactivated vaccine, but not in those inoculated with an mRNA vaccine

As aforementioned, the vaccines failed to constrain the duration of viral RNA shedding in mild COVID-19 patients (Figure 1C), which implies that vaccine induced immune responses might not play a pivotal role in determining viral persistence in these patients. To identify physiological variables that are relevant to the duration of viral RNA shedding in the mild COVID-19 patients, we performed correlation analyses between the duration of viral RNA shedding and 49 laboratory variables detected at the immediate early stage of infection. Our data showed that 16 variables in unvaccinated patients (Figure 2A) and 15 variables in patients inoculated with an inactivated vaccine (Figure 2B) correlated with the duration of viral RNA shedding, respectively. Of note, 10 variables were shared by these two groups of mild patients, including 3 positively correlated variables (Fibrinogen, Monocyte and IL-17) and 7 negatively correlated variables (Neutrophil, Eosinophil, Basophil, CD4, CD8, CD19 and CD16CD56 cell counts). In contrast, in patients inoculated with an mRNA vaccine, only eGFR (estimated glomerular filtering rate) was found to correlate negatively with the duration of viral RNA shedding (Figure 2C). Next, we compared the laboratory variables among the three groups of mild COVID-19 patients and found that inoculations with an inactivated vaccine or an mRNA vaccine influenced the laboratory parameters differentially (Supplemental table 1). Patients inoculated with an mRNA vaccine mounted highest median IL-8 and IL-17 responses at the immediate early stage of infection (Supplemental table 1).

**Figure 2.**
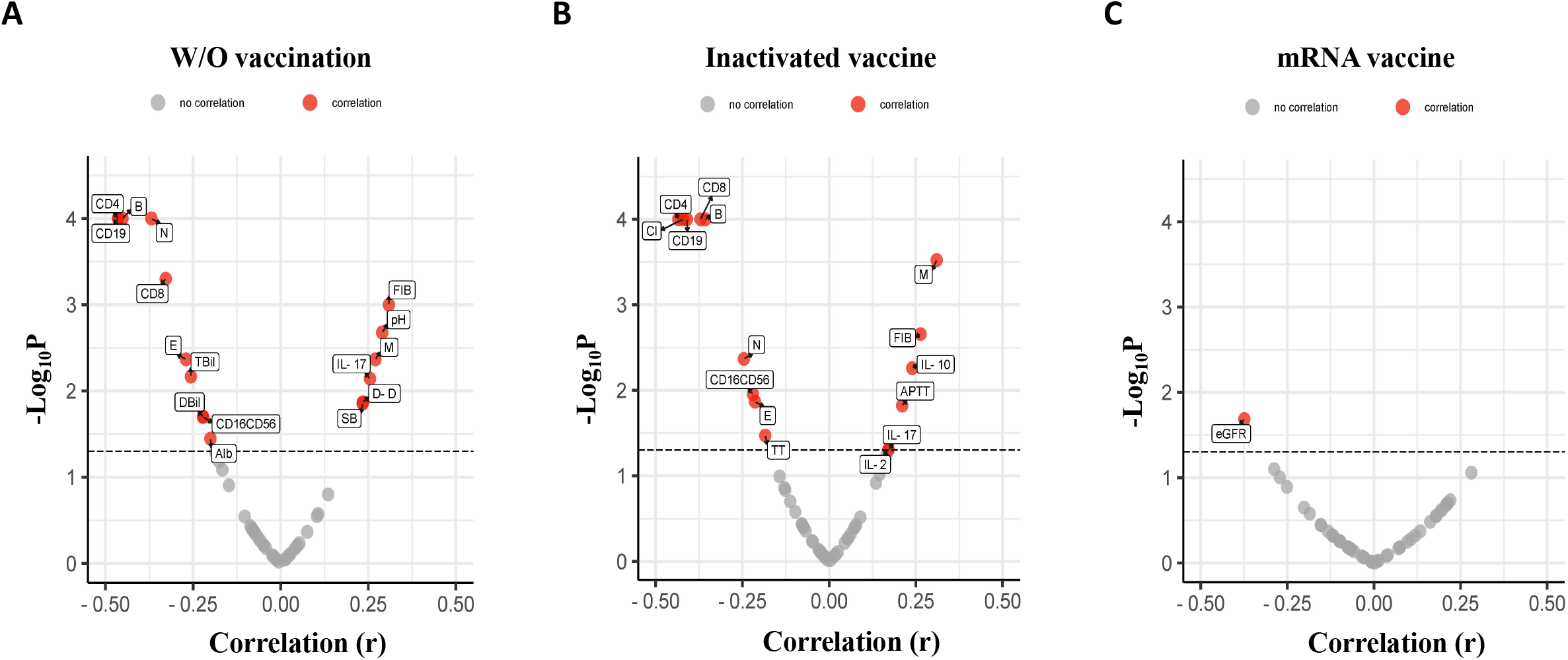
Correlation analyses between multiple laboratory variables and the duration of viral RNA shedding in different groups of mild COVID-19 patients. Data of 49 laboratory variables tested at the immediate early stage of infection were retrieved and correlations between each individual variable and the duration of viral RNA shedding were analyzed in unvaccinated mild patients (**A**), and mild patients inoculated with either an inactivated vaccine (**B**) or an mRNA vaccine (**C**), respectively. Variables that significantly correlated the duration of viral shedding (P < 0.05) were shown in red. Correlation analyses were done using Pearson correlation for normally distributed data and Spearman correlation for non-normally distributed data. Volcano plots were constructed using an online software (http://www.bioinformatics.com.cn).

### Variables correlating with the duration of viral RNA shedding in unvaccinated mild patients could predict the duration of viral RNA shedding in all groups of patients

Although multiple individual parameters were found to correlate significantly with the duration of viral RNA shedding, the correlation coefficients were low (│r│<0.5) (Figure 2), suggesting that the correlations were relatively weak. This notion was supported by the results of univariate logistic regression analyses, which showed that no individual variable could reliably discriminate between short duration of viral RNA shedding (≤14 days) from long shedding duration (>14 days) in unvaccinated mild patients (AUC<0.80) (Supplemental figure 2). The count of CD19^+^ B cell is of the highest discriminating efficiency (AUC=0.7556) followed by the count of CD4^+^ T cell (AUC=0.7261) (Supplemental figure 2). To further characterize the relationships between host factors and viral persistence, we performed multivariate logistic regression analyses leveraging the significant factors identified in the correlation analyses for unvaccinated mild patients. Our results showed that the selected laboratory variables discriminated between the short (≤14 days) and long (>14 days) viral RNA shedding duration quite well for all the three groups of mild COVID-19 patients. The area under the ROC curve (AUC) for unvaccinated patients, patients inoculated with an inactivated vaccine and patients inoculated with an mRNA vaccine were 0.9231, 0.8365 and 0.9508, respectively (Figure 3). Moreover, we found that this multivariate regression model could also predict the duration of viral RNA shedding in moderate COVID-19 patients. The AUC for unvaccinated moderate patients and moderate patients inoculated with an inactivated vaccine were 0.9110 and 0.9514, respectively (Figure 4). Due the limited sample size (only three patients), the multivariate regression analysis could not be applied to moderate patients inoculated with an mRNA vaccine.

**Figure 3.**
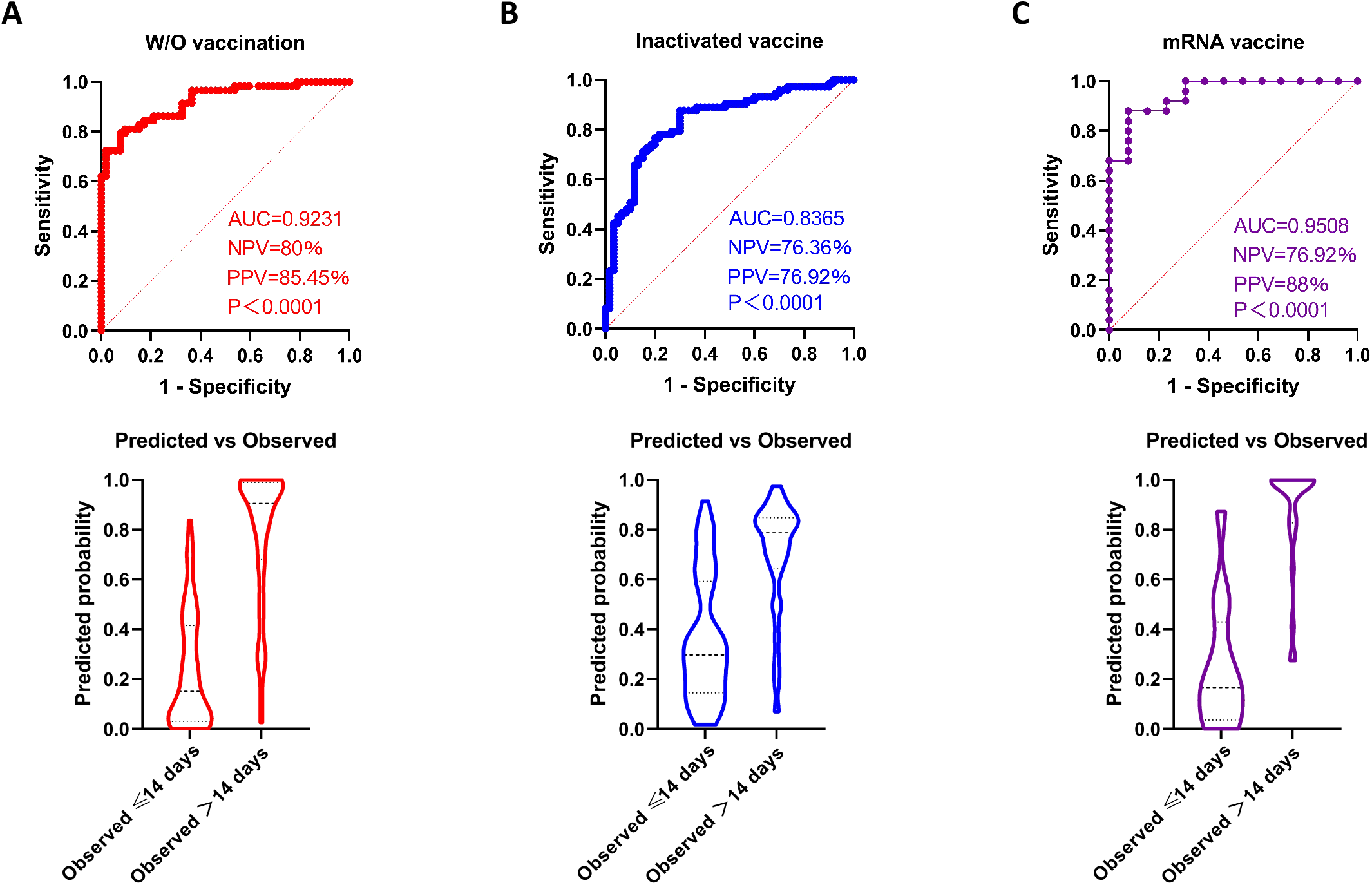
Variables correlating with viral persistence in unvaccinated mild patients could discriminate between short (≤14 days) and long (>14 days) duration of viral RNA shedding for all mild COVID-19 patients. Multivariate regression model constructed using significant factors identified in unvaccinated mild patients discriminated between short and long duration of viral RNA shedding for unvaccinated mild patients (**A**) and mild patients inoculated with an inactivated vaccine (**B**) or an mRNA vaccine (**C**). NPV, negative predictive value; PPV, positive predictive value.

**Figure 4.**
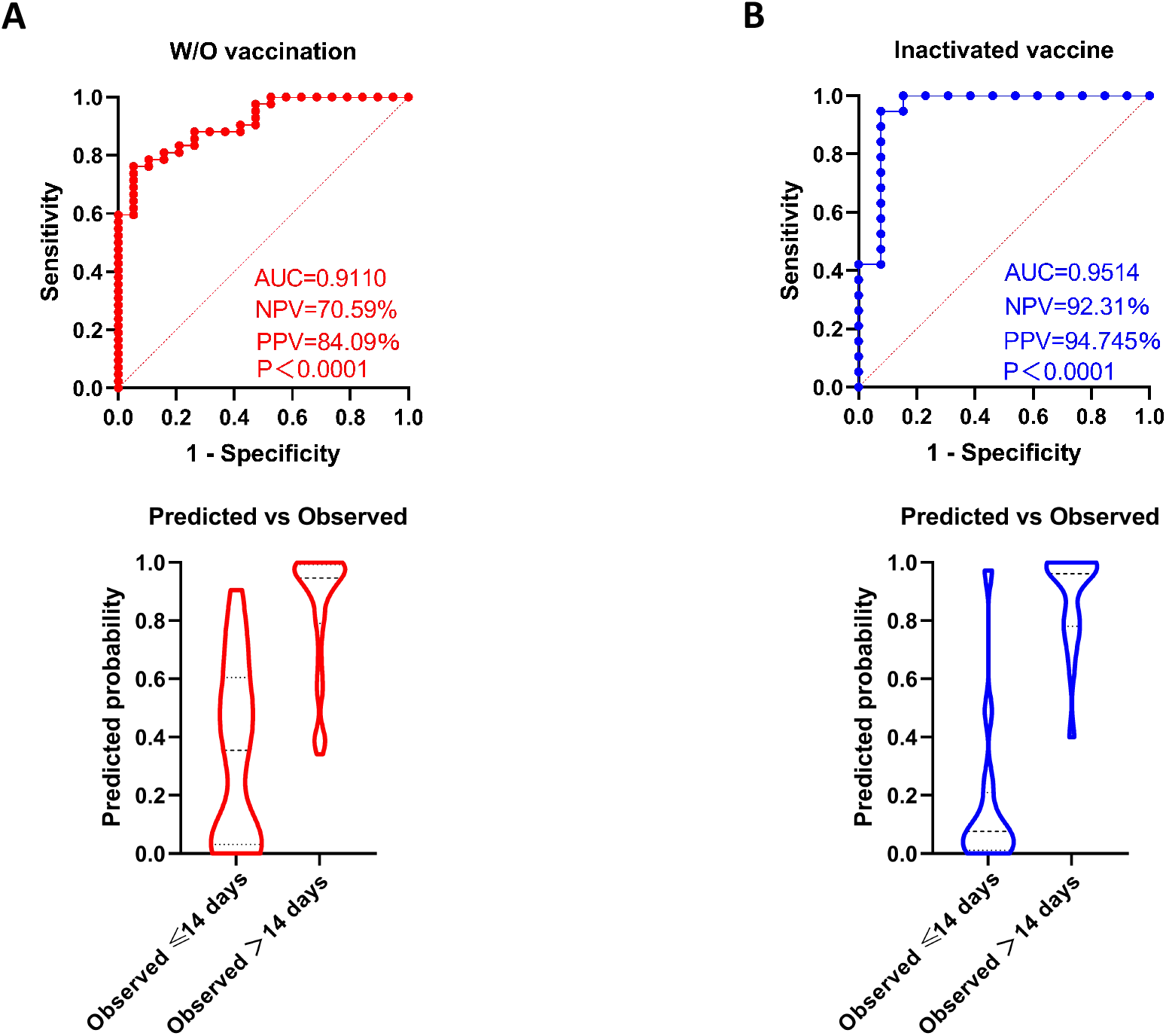
Variables correlating with viral persistence in unvaccinated mild patients could discriminate between short (≤14 days) and long (>14 days) duration of viral RNA shedding for moderate COVID-19 patients. Multivariate regression model constructed using significant factors identified in unvaccinated mild patients discriminated between short and long duration of viral RNA shedding for unvaccinated moderate patients (**A**) and moderate patients inoculated with an inactivated patients (**B**).

## Discussion

An important factor that influences the transmission of SARS-CoV-2 is the period of communicability, which may be influenced by many factors. To identify these factors, detections of live virus and viral RNA are frequently used as indicators of communicability, of which the live virus culture is thought to be more relevant to the communicability of SARS-CoV-2 infections (15-17).

However, the culture of live viruses is time-consuming and might be affected by specimen quality and the sensitivities of laboratory methods. Hence, as an alternative solution, the measurement of viral RNA shedding remains to be an important criterion for the diagnosis of COVID-19 and the discharge of hospitalized patients (18).

Multiple factors have been found to associate with prolonged viral RNA shedding in previous studies, such as old age (19), compromised immune status (20), treatment with corticosteroids (21-23) and disease severity (24, 25). Meanwhile, biological sex and comorbidities, such as hypertension and diabetes, were found not to associate with the viral RNA shedding (26-28). Unlike most previous works, our current study investigated the impacts of vaccination status and multiple laboratory variables in early infection on the duration of viral RNA shedding.

First of all, we compared the ratios of moderate COVID-19 cases among unvaccinated patients and patients fully vaccinated with either an inactivated vaccine or an mRNA vaccine. Our data showed that both the inactivated and the mRNA vaccines significantly reduced the incidence of moderate cases, which is in consistence with the observation of a previous real-world study (29). Next, we compared the duration of viral RNA shedding between unvaccinated and fully vaccinated patients. Quite unexpectedly, our results showed that vaccination significantly shortened the duration of virus shedding in moderate patients but not in mild patients, suggesting that vaccine induced specific immune responses did not contribute significantly to constraining viral persistence in mild patients. We were not able to elaborate the underlying mechanism in this retrospective study. However, as previous studies suggested that more severe COVID-19 could usually evoke stronger host immune responses (30, 31), we speculate that the memory immune responses established by vaccination could be more efficiently activated in moderate patients, which may control the in vivo virus replication better.

Next, to characterize host factors that may affect viral persistence in mild COVID-19 patients, we retrieved 49 laboratory variables which were detected at the immediate early stage after infection (at a median of 3 days post diagnosis). Correlation analyses showed that the plasma levels of IL-17, fibrinogen and the counts of peripheral leucocytes were associated with the duration of viral RNA shedding in both unvaccinated patients and patients inoculated with an inactivated vaccine. Elevated levels of proinflammatory cytokines and lymphopenia have been demonstrated to be associated with prolonged viral RNA shedding by previous studies (32-34). Here, we further showed that the counts of T, B, NK, neutrophil and eosinophil were negatively associated with the duration of viral RNA shedding, while the count of monocyte was positively associated with the duration of viral RNA shedding. More intriguingly, our data showed that all the significant individual variables identified in unvaccinated patients and patients inoculated with an inactivated vaccine were not significantly associated with the duration of viral RNA shedding in patients inoculated with an mRNA vaccine. This phenomenon implies that the mRNA vaccine may have unique impacts on host responses after break through infection. And these impacts might be deleterious, because the median duration of viral RNA shedding of patients inoculated with an mRNA vaccine tended to be longer than those of unvaccinated patients and patients inoculated with an inactivated vaccine. Insight into the underlying mechanisms will help to improve our understanding of the biological effect of mRNA vaccine.

At the end of this study, we established a multivariate regression model using the individual factors correlating with the duration of viral RNA shedding in unvaccinated mild patients and found that the model could be applied to all groups of patients, including the mild patients inoculated with an mRNA vaccine and the moderate patients. This finding suggests that despite of the impacts of vaccination status and the difference of disease severity, the combined effect of host factors on viral persistence remains stable.

Several limitations of the present study should be noted. First, antigen-specific immune responses were not detected in this retrospective work. Nonetheless, we speculate that the vaccine induce immunities might not be able to constrain viral RNA shedding in mild COVID-19 patients, since both an inactivated vaccine and an mRNA vaccine failed to shorten the duration of viral RNA shedding in these patients. Second, although a previous study showed that viral RNA load (>10^7^ copies/ml) is an independent risk factor for live virus shedding (35), the detection viral RNA might not always indicate the presence of live viruses. Further investigations are necessitated to characterize host factors that associate with the shedding of viable viruses. Third, most cases enrolled in this study were infected by either the D614G variant or the Delta variant, but we were not able to define the infected variant for each case. Therefore, the impact of viral factor on the duration of viral RNA shedding is out-of-scope for this study.

Despite of these limitations, our study showed for the first time that COVID-19 vaccines contributed to the clearance viral RNA in moderate cases, while failed to shorten the duration of viral RNA shedding in mild patients. Moreover, we identified a set of laboratory variables in early infection that could predict the persistence of viral RNA. These findings may serve as a rationale to optimize the control measures for COVID-19 pandemic.

## Data Availability

All data produced in the present work are contained in the manuscript

## Author contributions

*All authors confirmed they have contributed to the intellectual content of this paper and have met the following 4 requirements: (a) significant contributions to the conception and design, acquisition of data, or analysis and interpretation of data; (b) drafting or revising the article for intellectual content; (c) final approval of the published article; and (d) agreement to be accountable for all aspects of the article thus ensuring that questions related to the accuracy or integrity of any part of the article are appropriately investigated and resolved*.

ZZ and YW designed and supervised the study; XT and YZ collected the data; YW, XT and YZ analyzed the data; XT and YZ drafted the manuscript; YW, ZZ and WZ revised the manuscript. The underlying data have been verified by FF, ZZ and YW.

## Authors’ Disclosures or Potential Conflicts of Interest

*Upon manuscript submission, all authors completed the author disclosure form*.

*Disclosures and/or potential conflicts of interest:*

## Employment or Leadership

None declared.

## Consultant or Advisory Role

None declared.

## Stock Ownership

None declared.

## Honoraria

None declared.

## Research Funding

This work was partly supported by a grant from the National Natural Science Foundation of China (81971559), a grant from the major project of Study on Pathogenesis and Epidemic Prevention Technology System (2021YFC2302500) by the Ministry of Science and Technology of China and a grant from the Science and Technology Commission of Shanghai Municipality (21NL2600100).

## Expert Testimony

None declared.

## Patents

None declared.

## Acknowledgments

We thank Dr. Liqiu Jia and Ms. Jing Wang for their help in retrieving the clinical records of the COVID-19 patients.

---

**Supplementary figure 1.**
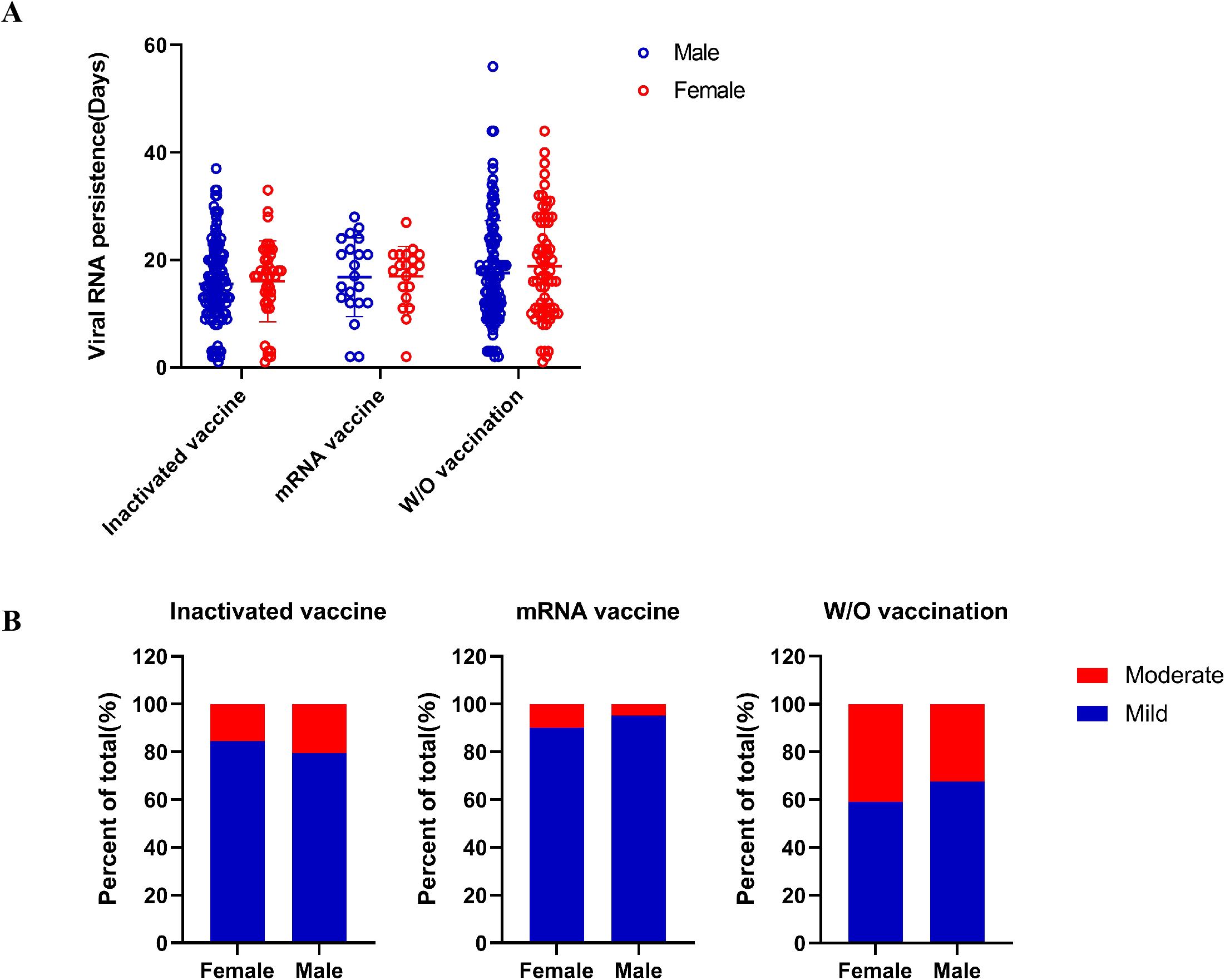
The average duration of viral RNA shedding and proportions of mild and moderate COVID-19 cases were similar between male and female patients. (A)The average duration of viral RNA shedding of male patients was similar with that of female patients. (B) The proportions of mild and moderate cases in male patients were comparable with those in female patients.

**Supplementary figure 2.**
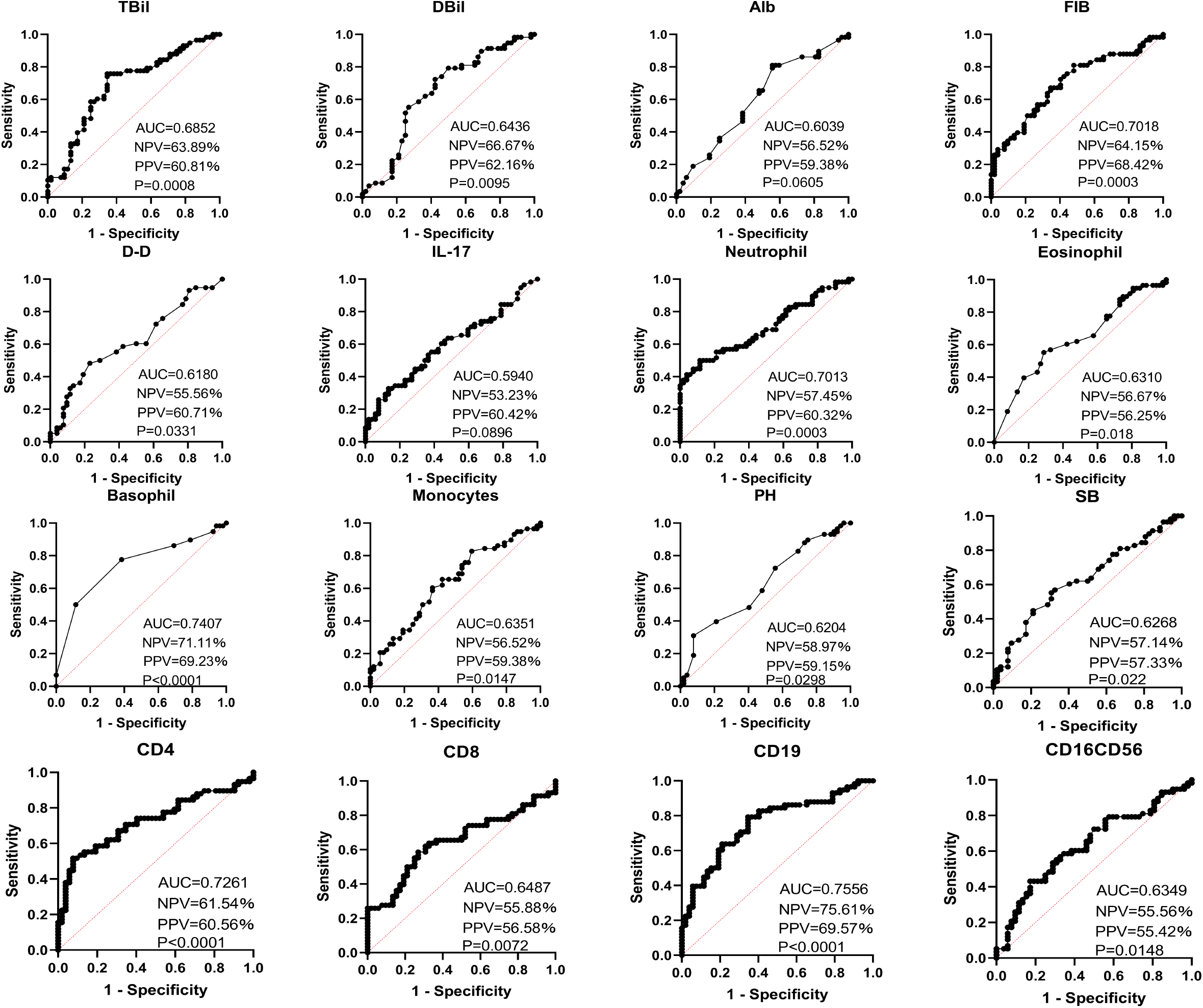
Individual variables could not reliably discriminate between short (≤14 days) and long (>14 days) duration of viral RNA shedding for unvaccinated mild COVID-19 patients. Univariate regression analyses showed that each of the significant variables identified in the correlation analyses could not confidently discriminate between short (≤14 days) and long (>14 days) duration of viral RNA shedding for the unvaccinated mild COVID-19 patients.

**Supplementary table 1.** Comparisons of demographical and laboratory variables among mild COVID-19 patients received the inactivated vaccine, the mRNA vaccine or no vaccine

| Variables | Normal range | (A)<br>Inactivated vaccine<br>(N=133) | (B)<br>mRNA vaccine<br>(N=38) | (C)<br>W/O vaccination<br>(N=110) | P value<br>(A) vs (C) | P value<br>(B) vs (C) | P value<br>(A) vs (B) |
| --- | --- | --- | --- | --- | --- | --- | --- |
| Age (years) | - | 34.0 (24.0-43.0) | 29.0 (25.0-44.5) | 31.0 (23-42.3) | 0.2228 | 0.7206 | 0.5906 |
| Gender (n) |  |  |  |  |  |  |  |
| Male | - | 100 | 20 | 71 | 0.0901 | 0.2461 | 0.0094 |
| Female | - | 33 | 18 | 39 |  |  |  |
| Time-since-vaccination<br>(Days) | - | 170.7±92.51 | 149.4±62.42 | - | - | - | 0.2778 |
| Duration of virus<br>shedding (Days) | - | 15 (11-20) | 17.5 (12-21) | 15 (9.75-23) | 0.8236 | 0.5911 | 0.6551 |
| Sampling timepoint<br>(Days post diagnosis) | - | 3 (2-3) | 3 (2-4) | 3 (2-3) | 0.9769 | 0.1001 | 0.0670 |
| Biochemistry |  |  |  |  |  |  |  |
| ALT (U/L) | 7-40 | 18 (13-27) | 16 (12-26.75) | 17.5 (12-28) | 0.6663 | 0.9068 | 0.6321 |
| AST (U/L) | 13-35 | 18 (15-23.5) | 18.5 (16.75-24) | 18 (16-21) | 0.5972 | 0.3879 | 0.6472 |
| ALP (U/L) | 35-100 | 67 (54.5-81.5) | 60.5 (55-70.5) | 65 (53-60) | 0.5150 | 0.4316 | 0.1434 |
| L-γ-GGT (U/L) | 7-45 | 20 (15-35) | 20.5 (13-33) | 20.5 (14-30.25) | 0.5125 | 0.5942 | 0.2686 |
| LDH (U/L) | 120-250 | 181 (159-205) | 190.6±48.38 | 177 (157.8-201) | 0.4230 | 0.3417 | 0.6914 |
| TBil (μmol/L) | 3.4-20.5 | 7.8 (5.65-11.1) | 6.5 (4.75-8.15) | 7.6 (5.675-10.48) | 0.7469 | 0.0229 | 0.0098 |
| DBil (μmol/L) | 0-8.6 | 4.1 (3.2-5.7) | 3.5 (2.975-4.3) | 3.7 (3-4.925) | 0.0254 | 0.4444 | 0.0155 |
| Alb (g/L) | 40-55 | 45 (43-47) | 44.24±2.765 | 44.89±3.237 | 0.7724 | 0.2695 | 0.1526 |
| Glb (g/L) | 20-40 | 30 (27-33) | 29.16±4.290 | 28.79±4.182 | 0.0123 | 0.6438 | 0.0962 |
| **A/G (%)** | 1.2-2.4 | 1.5 (1.375-1.655) | 1.546±0.2328 | 1.591±0.2536 | 0.0383 | 0.3409 | 0.5021 |
| **K (mmol/L)** | 3.5-5.3 | 3.875±0.2996 | 3.961±0.3796 | 3.9 (3.7-4.125) | 0.0758 | 0.8492 | 0.1475 |
| **Na (mmol/L)** | 137-147 | 140.2±1.885 | 141 (140-142) | 139.9±1.812 | 0.1460 | 0.0042 | 0.0896 |
| **Cl (mmol/L)** | 99-110 | 104.3±1.946 | 104.3±1.927 | 104.7±2.379 | 0.2555 | 0.3616 | 0.8335 |
| **Urea (mmol/L)** | 2.6-7.5 | 4 (3.3-4.65) | 3.766±0.8438 | 3.98 (3.4-4.605) | 0.9671 | 0.0619 | 0.0729 |
| **SCr (μmol/L)** | 41-73 | 76.07±15.62 | 70.22±13.58 | 70.87±13.49 | 0.0065 | 0.7999 | 0.0379 |
| **Glu (mmol/L)** | 3.9-6.1 | 5.77 (5.14-6.38) | 5.565 (5.208-6.218) | 5.535 (4.948-6.553) | 0.4672 | 0.5734 | 0.8831 |
| **UA (μmol/L)** | 150-350 | 342.8±91.94 | 313.1±98.38 | 333.1±86.62 | 0.4024 | 0.2392 | 0.0862 |
| **eGFR (mL/ (min\*1.73m²))** | >90 | 104.2±17.67 | 104.2 (95.29-113.3) | 111.1±19.51 | 0.0044 | 0.1141 | 0.4159 |
| **Mb(ng/mL)** | <25 | 7.890 (5.325-10.32) | 8.620 (6.210-13.25) | 8.495 (5.555-11.97) | 0.2868 | 0.6174 | 0.2159 |
| **NT-proBNP (pg/mL)** | 0-500 | 69.30 (61.45-181.5) | 64.65 (60.83-96.38) | 70.51 (60.7-178.9) | 0.8380 | 0.2436 | 0.1946 |
| **CK Isoenzyme (ng/mL)** | <5 | 0.46 (0.3-1.1) | 0.7195±0.5253 | 1.21 (0.6-1.82) | <0.0001 | 0.0002 | 0.4459 |
| **Blood coagulation** |  |  |  |  |  |  |  |
| **INR** | 0.8-1.3 | 0.94 (0.91-0.98) | 0.9368±0.0599 | 0.96 (0.91-1.013) | 0.1000 | 0.0774 | 0.5193 |
| **PTA (%)** | 75-100 | 111.5±13.62 | 109.5 (105-122.5) | 108 (96-118.3) | 0.0623 | 0.0803 | 0.5817 |
| **PT(S)** | 11-14.3 | 12.6 (12.2-13) | 12.52±0.6475 | 12.8 (12.3-13.43) | 0.0348 | 0.0157 | 0.3332 |
| **APTT(S)** | 32-43 | 39.56±4.245 | 40.38±4.284 | 40 (37.43-43.30) | 0.5022 | 0.1611 | 0.2979 |
| **FIB (g/L)** | 2-4 | 3.097±0.6709 | 3.31 (2.875-3.95) | 2.88 (2.553-3.258) | 0.0555 | 0.0003 | 0.0134 |
| **FDP (μg/mL)** | 0-5 | 1.5 (1.13-2.185) | 1.295 (0.9975-1.615) | 1.620 (0.6425-3.130) | 0.7372 | 0.2508 | 0.0246 |
| **D-D (μg/mL)** | 0-0.5 | 0.2000 (0.16-0.295) | 0.2100 (0.18-0.235) | 0.2200 (0.17-0.2725) | 0.6326 | 0.5744 | 0.9049 |
| **TT (S)** | 14-21 | 16 (15.35-16.95) | 15.6 (14.68-16) | 16.15 (15.5-16.6) | 0.7475 | 0.0003 | 0.0023 |
| **ESR (mm/h)** | 0-20 | 7 (4-13) | 8.5 (4-19.75) | 10 (5-22) | 0.0008 | 0.2571 | 0.2900 |
| **Cytokine** |  |  |  |  |  |  |  |
| **IL-2 (pg/mL)** | <7.5 | 1.38 (0.27-3.41) | 2.19 (0.52-4.118) | 0.235 (0-0.9725) | <0.0001 | <0.0001 | 0.1063 |
| IL-8 (pg/mL) | <20.6 | 12.77 (0-60.27) | 55.5 (0-165.5) | 0.115 (0-2.01) | <0.0001 | <0.0001 | 0.0286 |
| IL-10 (pg/mL) | <12.9 | 1.51 (0.42-2.475) | 1.135 (0.4475-2.488) | 0.78 (0.3125-1.623) | 0.0005 | 0.0627 | 0.6751 |
| IL-17 (pg/mL) | <21.4 | 3.52 (1.48-56.29) | 57.68 (2.698-71.63) | 1.065 (0.1925-2.505) | 0.0005 | <0.0001 | 0.0016 |
| **Leucocyte count** |  |  |  |  |  |  |  |
| N (×10<sup>9</sup>/L) | 1.8-6.3 | 3.59 (2.7-4.675) | 3.848±1.379 | 3.828±1.486 | 0.7770 | 0.9411 | 0.7468 |
| E (×10<sup>9</sup>/L) | 0.02-0.52 | 0.070 (0.03-0.13) | 0.070 (0.02-0.1525) | 0.070 (0.02-0.13) | 0.6821 | 0.5316 | 0.7789 |
| B (×10<sup>9</sup>/L) | 0-0.06 | 0.0200 (0.01-0.03) | 0.0200 (0.01-0.03) | 0.0200 (0.01-0.03) | 0.9393 | 0.5578 | 0.5792 |
| M (×10<sup>9</sup>/L) | 0.1-0.6 | 0.57 (0.435-0.705) | 0.525 (0.438-0.745) | 0.54 (0.42-0.66) | 0.3459 | 0.3953 | 0.8106 |
| CD4 (cell/μL) | 410-1590 | 496.5 (365.1-735.5) | 582.4±235 | 600.5 (404.5-824.1) | 0.0601 | 0.4985 | 0.5421 |
| CD8 (cell/μL) | 190-1140 | 388.4 (239.6-526.1) | 381.0 (285.6-492.3) | 371.6 (257.4-583.5) | 0.4456 | 0.8143 | 0.7625 |
| CD19 (cell/μL) | 107-698 | 152.3 (99.97-267.2) | 187±92.56 | 193.7 (111.3-303.9) | 0.0218 | 0.2723 | 0.5324 |
| CD16CD56 (cell/μL) | 95-640 | 254.0 (166-356) | 276.7±158.7 | 229.0 (145-342.5) | 0.2926 | 0.5310 | 0.9256 |
| CD4/CD8 | 0.9-3.6 | 1.480 (1.12-1.85) | 1.350 (1.088-1.633) | 1.452 (1.167-1.86) | 0.7462 | 0.2349 | 0.3016 |
| **Arterial blood gas** |  |  |  |  |  |  |  |
| pH | 7.35-7.45 | 7.37 (7.34-7.39) | 7.36 (7.348-7.38) | 7.38 (7.36-7.4) | 0.0694 | 0.0230 | 0.3442 |
| pCO<sub>2</sub> (KPa) | 4.3-6 | 5.952±0.6798 | 6.005 (5.718-6.323) | 5.685±0.5314 | 0.0009 | 0.0005 | 0.4094 |
| pO<sub>2</sub> (KPa) | 10-14 | 13.60 (12.2-14.9) | 14.04±2.162 | 13.45 (12.48-15.03) | 0.9175 | 0.3166 | 0.2090 |
| SB (mmol/L) | 21.3-24.8 | 24.30 (23.2-25) | 24.28±1.030 | 24.10 (23.1-24.83) | 0.2988 | 0.1956 | 0.5894 |
| SpO<sub>2</sub>(%) | 91.9-99 | 98.10 (97.15-98.75) | 98.35 (97.85-98.8) | 98.10 (97.4-98.7) | 0.4967 | 0.3560 | 0.1641 |
| AB (mmol/L) | 22-29 | 25.43±1.825 | 25.8 (25.03-26.7) | 25 (24.08-25.8) | 0.0105 | 0.0036 | 0.4427 |

**Note:** Data conforming to the normal distribution are shown as mean ± standard deviation (SD), and data not conforming to the normal distribution are shown as the median (IQR).
**Abbreviations:** ALT, glutamic pyruvic transaminase; AST, glutamic oxalo-acetic transaminase; ALP, alkaline phosphatase; L-γ-GGT, gamma glutamyl transferase; LDH, lactate dehydrogenase; TBil, total bilirubin; DBil, Direct Bilirubin; Alb, albumin; globulin; SCr, serum creatinine; Glu, glucose; UA, uric acid; eGFR, estimated glomerular filtering rate; INR, international normalized ratio; PTA, prothrombin activity prothrombin time activity; PT, Prothrombin
time; APTT, activated partial thromboplastin time; FIB, Fibrinogen; FDP, fibrin/fibrinogen degradation products; D-D, D-dimer; TT, thrombin time; Mb, myoglobin; NT-proBNP, N terminal pro B type natriuretic peptide; CK Isoenzyme, creatine kinase isoenzyme; ESR, erythrocyte sedimentation rate; N, neutrophil; E, eosinophil; B, basophil; M, monocyte; SB, standard bicarbonate; AB, actual bicarbonate.

## References

1. Liu Q, Qin C, Liu M, Liu J. Effectiveness and safety of SARS-CoV-2 vaccine in real-world studies: a systematic review and meta-analysis. Infect Dis Poverty 2021;10:132.

2. Zheng C, Shao W, Chen X, Zhang B, Wang G, Zhang W. Real-world effectiveness of COVID-19 vaccines: a literature review and meta-analysis. Int J Infect Dis 2022;114:252–60.

3. Hungerford D, Cunliffe NA. Real world effectiveness of covid-19 vaccines. 2021;374:n2034.

4. Shah ASV, Gribben C, Bishop J, Hanlon P, Caldwell D, Wood R, et al. Effect of Vaccination on Transmission of SARS-CoV-2. N Engl J Med 2021;385:1718–20.

5. Harris RJ, Hall JA, Zaidi A, Andrews NJ, Dunbar JK, Dabrera G. Effect of Vaccination on Household Transmission of SARS-CoV-2 in England. N Engl J Med 2021;385:759–60.

6. Eyre DW, Taylor D, Purver M, Chapman D, Fowler T, Pouwels KB, et al. The impact of SARS-CoV-2 vaccination on Alpha & Delta variant transmission. Preprint at https://www.medrxiv.org/content/10.1101/2021.09.28.21264260v1 (2021).

7. Vitiello A, Ferrara F, Troiano V, La Porta R. COVID-19 vaccines and decreased transmission of SARS-CoV-2. Inflammopharmacology 2021;29:1357–60.

8. Callaway E. Delta coronavirus variant: scientists brace for impact. Nature 2021;595:17–8.

9. Levine-Tiefenbrun M, Yelin I, Katz R, Herzel E, Golan Z, Schreiber L, et al. Initial report of decreased SARS-CoV-2 viral load after inoculation with the BNT162b2 vaccine. Nat Med 2021;27:790–2.

10. Levine-Tiefenbrun M, Yelin I, Alapi H, Katz R, Herzel E, Kuint J, et al. Viral loads of Delta-variant SARS-CoV-2 breakthrough infections after vaccination and booster with BNT162b2. Nat Med 2021;27:2108–10.

11. Pouwels KB, Pritchard E, Matthews PC, Stoesser N, Eyre DW, Vihta K-D, et al. Impact of Delta on viral burden and vaccine effectiveness against new SARS-CoV-2 infections in the UK. Preprint at https://www.medrxiv.org/content/10.1101/2021.08.18.21262237v1 (2021).

12. Riemersma KK, Grogan BE, Kita-Yarbro A, Halfmann PJ, Segaloff HE, Kocharian A, et al. Shedding of Infectious SARS-CoV-2 Despite Vaccination. Preprint at https://www.medrxiv.org/content/10.1101/2021.07.31.21261387v2 (2021).

13. Salvatore PP, Lee CC, Sleweon S, McCormick DW, Nicolae L, Knipe K, et al. Transmission potential of vaccinated and unvaccinated persons infected with the SARS-CoV-2 Delta variant in a federal prison, July—August 2021. Preprint at https://www.medrxiv.org/content/10.1101/2021.11.12.21265796v1 (2021).

14. Diagnosis and Treatment Protocol for Novel Coronavirus Pneumonia (Trial Version 7). Chin Med J 2020;133:1087–95.

15. Manzulli V, Scioscia G, Giganti G, Capobianchi MR, Lacedonia D, Pace L, et al. Real Time PCR and Culture-Based Virus Isolation Test in Clinically Recovered Patients: Is the Subject Still Infectious for SARS-CoV2? J Clin Med 2021;10:309.

16. Huang CG, Lee KM, Hsiao MJ, Yang SL, Huang PN, Gong YN, et al. Culture-Based Virus Isolation To Evaluate Potential Infectivity of Clinical Specimens Tested for COVID-19. J Clin Microbiol 2020;58:e01068–e20.

17. Jefferson T, Spencer EA, Brassey J, Heneghan C. Viral Cultures for Coronavirus Disease 2019 Infectivity Assessment: A Systematic Review. Clin Infect Dis 2021;73:e3884–e99.

18. Jin YH, Zhan QY, Peng ZY, Ren XQ, Yin XT, Cai L, et al. Chemoprophylaxis, diagnosis, treatments, and discharge management of COVID-19: An evidence-based clinical practice guideline (updated version). Mil Med Res 2020;7:41.

19. Cevik M, Tate M, Lloyd O, Maraolo AE, Schafers J, Ho A. SARS-CoV-2, SARS-CoV, and MERS-CoV viral load dynamics, duration of viral shedding, and infectiousness: a systematic review and meta-analysis. Lancet Microbe 2021;2:e13–e22.

20. Zhu L, Gong N, Liu B, Lu X, Chen D, Chen S, et al. Coronavirus Disease 2019 Pneumonia in Immunosuppressed Renal Transplant Recipients: A Summary of 10 Confirmed Cases in Wuhan, China. Eur Urol 2020;77:748–54.

21. Hu F, Yin G, Chen Y, Song J, Ye M, Liu J, et al. Corticosteroid, oseltamivir and delayed admission are independent risk factors for prolonged viral shedding in patients with Coronavirus Disease 2019. Clin Respir J 2020;14:1067–75.

22. Liu W, Liu Y, Xu Z, Jiang T, Kang Y, Zhu G, et al. Clinical characteristics and predictors of the duration of SARS-CoV-2 viral shedding in 140 healthcare workers. J Intern Med 2020;288:725–36.

23. Qi L, Yang Y, Jiang D, Tu C, Wan L, Chen X, et al. Factors associated with the duration of viral shedding in adults with COVID-19 outside of Wuhan, China: a retrospective cohort study. Int J Infect Dis 2020;96:531–7.

24. Park M, Pawliuk C, Nguyen T, Griffitt A, Dix-Cooper L, Fourik N, et al. Determining the period of communicability of SARS-CoV-2: A rapid review of the literature. Preprint at https://www.medrxiv.org/content/10.1101/2020.07.28.20163873v1

25. Fontana LM, Villamagna AH, Sikka MK, McGregor JC. Understanding viral shedding of severe acute respiratory coronavirus virus 2 (SARS-CoV-2): Review of current literature. Infect Control Hosp Epidemiol 2021;42:659–68.

26. Stehlik P, Alcorn K, Jones A, Schlebusch S, Wattiaux A, Henry DA. Repeat testing for SARS-CoV-2: persistence of viral RNA is common, and clearance is slower in older people. Med J Aust 2021;214:468–70.

27. Xu K, Chen Y, Yuan J, Yi P, Ding C, Wu W, et al. Factors Associated With Prolonged Viral RNA Shedding in Patients with Coronavirus Disease 2019 (COVID-19). Clin Infect Dis 2020;71:799–806.

28. Zhou C, Zhang T, Ren H, Sun S, Yu X, Sheng J, et al. Impact of age on duration of viral RNA shedding in patients with COVID-19. Aging (Albany NY) 2020;12:22399–404.

29. Tenforde MW, Self WH, Adams K, Gaglani M, Ginde AA, McNeal T, et al. Association Between mRNA Vaccination and COVID-19 Hospitalization and Disease Severity. Jama 2021;326:2043–54.

30. Mariën J, Ceulemans A, Michiels J, Heyndrickx L, Kerkhof K, Foque N, et al. Evaluating SARS-CoV-2 spike and nucleocapsid proteins as targets for antibody detection in severe and mild COVID-19 cases using a Luminex bead-based assay. J Virol Methods 2021;288:114025.

31. Reynolds CJ, Swadling L, Gibbons JM, Pade C, Jensen MP, Diniz MO, et al. Discordant neutralizing antibody and T cell responses in asymptomatic and mild SARS-CoV-2 infection. Sci Immunol 2020;5:eabf3698.

32. Gao C, Zhu L, Jin CC, Tong YX, Xiao AT, Zhang S. Proinflammatory cytokines are associated with prolonged viral RNA shedding in COVID-19 patients. Clin Immunol 2020;221:108611.

33. Hao S, Lian J, Lu Y, Jia H, Hu J, Yu G, et al. Decreased B Cells on Admission Associated With Prolonged Viral RNA Shedding From the Respiratory Tract in Coronavirus Disease 2019: A Case-Control Study. J Infect Dis 2020;222:367–71.

34. Mondi A, Lorenzini P, Castilletti C, Gagliardini R, Lalle E, Corpolongo A, et al. Risk and predictive factors of prolonged viral RNA shedding in upper respiratory specimens in a large cohort of COVID-19 patients admitted to an Italian reference hospital. Int J Infect Dis 2021;105:532–9.

35. van Kampen JJA, van de Vijver D, Fraaij PLA, Haagmans BL, Lamers MM, Okba N, et al. Duration and key determinants of infectious virus shedding in hospitalized patients with coronavirus disease-2019 (COVID-19). Nat Commun 2021;12:267.

